# “Just Saiyan: Tail-trimming Human Monkeypox Virus Assemblies Emphasizes Resolvable Regions in Inverted Terminal Repeats to Improve the Resolution of Reference and Production Genomes for Genomic Surveillance”

**DOI:** 10.1101/2022.09.06.22279633

**Authors:** Alejandro R. Gener

## Abstract

Our ability to track the spread of the human monkeypox virus (hMPXV) during the ongoing monkeypox (hMPX) outbreak of 2022 relies on the availability of high-quality reference genomes. However, the way the information content of these genomes is organized in genome databases leaves room for interpretation. A current limitation of hMPXV genomic analysis is that the variability occurring in the inverted terminal repeats (ITRs) cannot be effectively resolved. This is because of shortcomings of the leading short-read sequencing and reference-guided assembly and variant calling used in the ongoing global hMPXV outbreak surveillance effort. Here I propose ITR tail-trimming, a simple no-cost reframing of how we organize hMPXV reference genomes and future assemblies. This approach is based on long terminal repeat (LTR) tail-trimming, which is a common practice in HIV sequence analysis. The main point of repeat sequence trimming is to remove problematic sequences while paying attention to limitations of mapping and variant calling in remaining repeat-associated (but ideally no longer repetitive) sequence. ITR tail-trimming would neutralize ITRs as distracting features at the read- and assembly-levels, allowing the global community to focus our efforts to track variability across hMPXV genomes.

## Introduction

The ongoing monkeypox (hMPX) **outbreak** of 2022 (WHO, 2022) coincided with unprecedented global genomic surveillance networks able to contribute to publicly available monkeypox virus (hMPXV) genome assemblies. Our ability to diagnose infection and to track the spread of this emerging pathogen relies on the availability of high-quality genomes. However, the way the information content of these genomes is organized in genome databases leaves room for interpretation. Specifically, sequence data of inverted terminal repeats (ITR) with effectively identical internal sequence are often not unambiguously callable (phasable) because sequencing reads are often much shorter than the 6.5kb∼ hMPXV ITRs (**Figure 1: hMPXV ITRs**).

**Figure 1:**
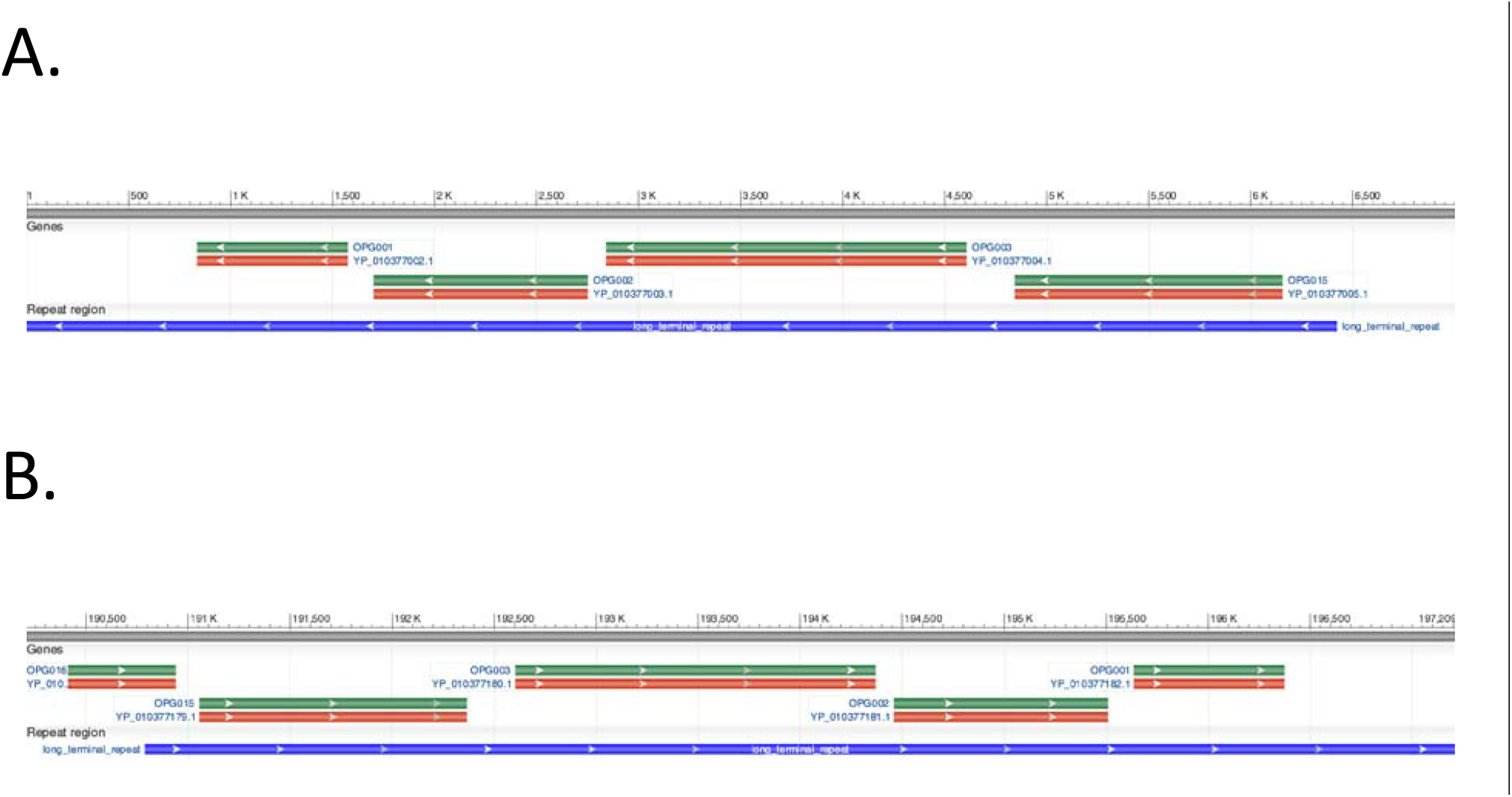
hMPXV ITRs. hMPXV 5’ (**1A**) and 3’ (**1B**) ITRs. Image from NCBI Graphics RefSeq:NC_063383.1. Each ITR is 6422 bp-long.

Reference genomes and production genomes for hMPXV are organized as non-ITR sequence sandwiched between two complete ITRs (e.g., 5’ITR-body-3’ITR). hMPXVs are closely related DNA viruses with relatively low mutation rates (10^−^ 5 to 10^−^ 6 per site) ((ViralZone, 2022); (Kerr et al., 2012)). For perspective, the current hMPXV outbreak’s reference genome RefSeq:NC_063383.1 (Clade IIb, formerly “West African” (Happi et al., 2022)) is 197,209 bases long. Early cases from the current 2022 hMPX outbreak had less than 10 acquired mutations compared to GenBank:MT903343 (Gigante et al., 2022). This is consistent with what was seen in an earlier 2017 hMPX outbreak in which authors noted mutation burdens as low as 0.4-1.5 single nucleotide polymorphisms per genome in a given transmission chain (Mauldin et al., 2022). Given the low mutation rate and low cluster divergences, care must be taken to rule out sequencing error to be able to assess genomes from suspected cluster cases.

The monkeypox-specific primers in use by the CDC are anchored in the ITRs (CDC Poxvirus & Rabies Branch (PRB), 2022). As of 9/2/2022, the current CDC-recommended MPXV-specific primers overlap the OPG002 gene. However, use of ITRs is risky as a sole hMPXV-specific target because of the potential for internal sequence heterogeneity to obscure ITR sequence and possibly structure (Gubser & Smith, 2002).

ITRs are also important in part because they harbor protein-coding genes. In order from the current assembly’s 5’ start site: OPG001, OPG002, OPG003, OPG015. The OPG001 gene produces MPXVgp001/ Chemokine binding protein (Cop-C23L) and is annotated in the Reference Sequence (RefSeq) record to be the most abundantly expressed secreted MPXV protein. The OPG002 gene product NBT03_gp002/ CrmB is a TNF-alpha-receptor-like protein. The OPG003 gene makes MPXVgp003/Ankyrin repeat protein (25). The OPG015 gene makes MPXVgp004/Ankyrin repeat protein (39).

In a recent presentation on June 22, 2022, Dr. Gustavo Palacios of Icahn School of Medicine at Mount Sinai suggested in passing that the hMPXV ITR could be either excluded from posterior analysis or treated as one copy. hMPXV ITRs may obscure mapping of short-reads depending on how internal repeats are arranged. Others noted that poxviruses can have repetitive low complexity regions in their ITRs (Gubser & Smith, 2002). These had been seen to some extent in hMPXV sequences, framed in the language of short tandem repeats (STR) in monkeypox viruses (Kugelman et al., 2014). An open question was whether these kinds of repetitive sequences occurred in hMPXV ITRs during the current outbreak, and whether these could interfere with mapping of the kinds of reads that were currently being used to help assemble hMPX genomes.

Regardless of STRs, variability within ITRs is currently obscured, or camouflaged (*Ebbert et al., 2019*), preventing simpler haploid/single-copy mapping and variant analysis. Their display as full-length inverted copies may be problematic depending on the sequencing platform used to make the assemblies. Nanopore (Oxford Nanopore Technologies; ONT) platforms are more flexible in their input and may receive as input direct/native DNA (PCR-free; not including multiplexing) or amplicon. On 9/4/22 there were no hMPXV assemblies made with PacBio instruments in NCBI. (Search term used: “(“Monkeypox virus”[Organism] OR “monkeypox virus”[All Fields]) AND pacbio[All Fields]”.) For higher-throughput Illumina (ILM) platforms (e.g., NovaSeq 6000), the chemistry is optimized for paired-end 150 base-long and an insert size (total DNA piece length) around 300 bases if using paired-end sequencing. Other common ILM platforms use PE150 and similar insert sizes too. So, identified variability beyond the 300-base ITR threshold is unlikely to be resolvable to either IT. In other words, variants within deeper “tails” are not phasable, though they may be identified in read data and may make it into assemblies depending on the allele frequency thresholds used to call variants.

Here I propose ITR tail-trimming, a simple no-cost reframing of how we organize hMPXV reference genomes and future assemblies which would neutralize ITRs as distracting features at the read- and assembly-levels, allowing the global community to focus our efforts to track variability across the hMPXV genome. This process has been employed implicitly for HIV RNA sequencing analyses and may benefit the hMXP/hMPXV research communities during the ongoing outbreak.

## Results and Discussion

### HIV LTRs are repeat sequences that set a precedent for trimming

Prior to the current hMPX outbreak and coronavirus pandemic, the most studied human pathogen was HIV-1. As such, insights from HIV-1 genomics may be transferable to other virus pathogen systems such as MPXV.

The HIV-1 genome was originally represented as a DNA provirus that was captured by restriction cloning into a molecular clone/plasmid (Gener et al., 2021). This enabled subsequent molecular biological dissection of the virus, which is still ongoing after 37+ years (Gener, 2022). Prominent structural features of HIV proviruses include long terminal repeats (LTRs) (**Figure 2A**) (Starcich et al., 1985) (Krebs et al., 2001).

**Figure 2:**
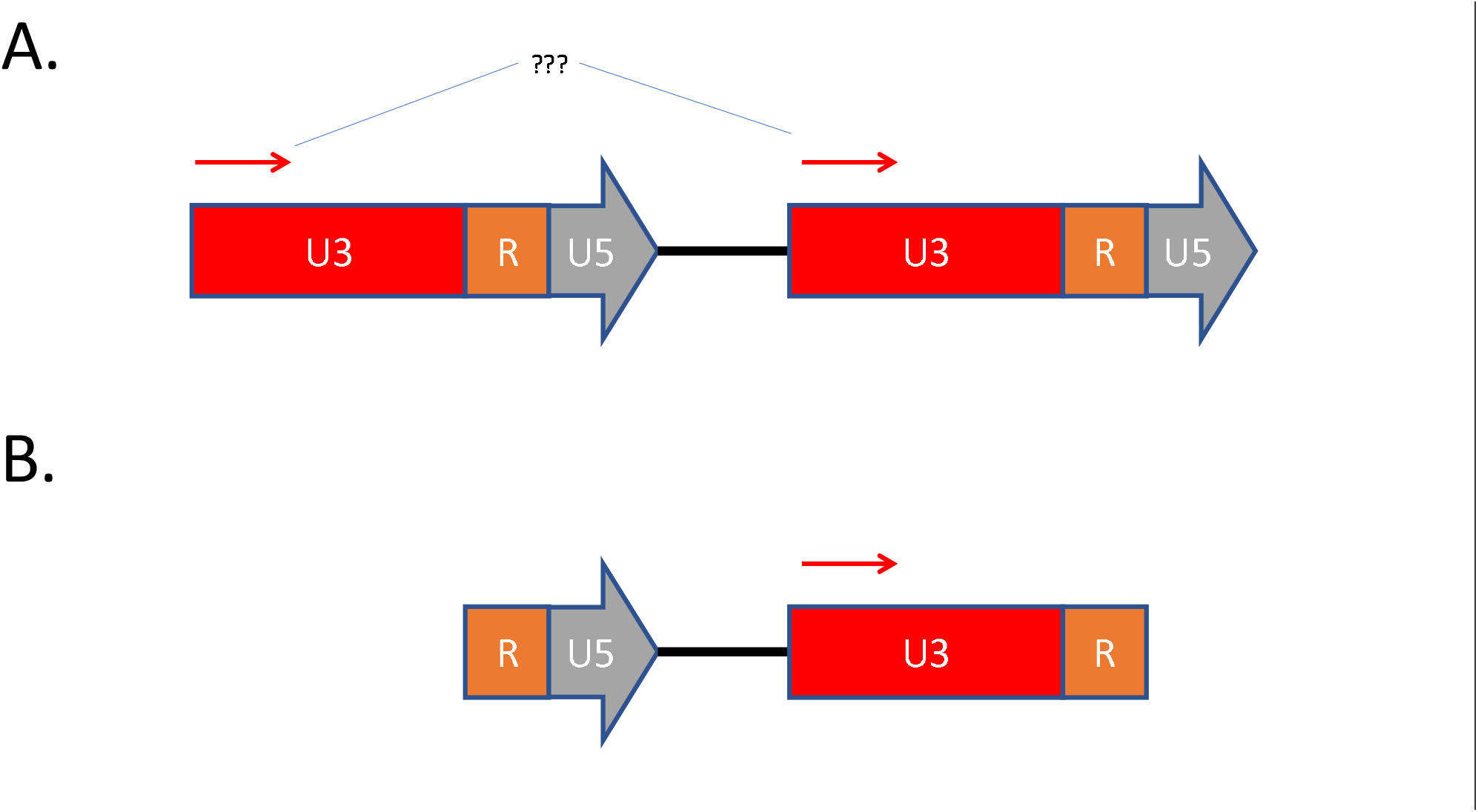
HIV LTR tail-trimming facilitates read mapping. The 5’ (upstream) LTR sequence is modeled to proceed in the forward direction, which is the same as the HIV open reading frames (not shown for simplicity). Figure not to scale. **2B**: The most common current way to model the HIV genome is to break up the LTR into 5’ R-U5 and 3’ U3-R segments. This avoids ambiguous mapping and simplifies analyses. Here, the “tail” of the 5’ LTR is trimmed because it can be represented as at the 3’ end; and the “arrow” 3’ U5 is trimmed. This region contains the main functional HIV polyA signal that is removed during host cell mRNA processing.

HIV LTRs are large (∼640 bp) proviral genomic features analogous to hMPXV ITRs. HIV LTRs occur in identical (arbitrarily forward) orientation, flank the 5’ (head) and 3’ (tail) ends of the full-length unspliced replication-competent provirus. These LTRs were longer than the many iterations of short-read sequencing, increasing from 50, 75, 150 (less commonly >250) single-end and 150 (less commonly >250) paired-end Illumina sequencing by synthesis (Shendure et al., 2017). Other sequencing methods like Sanger sequencing or ion torrent, or newer ILM methods could increase read lengths to nearly LTR-length reads. However, the LTR lengths and additional sequence complexity represent a current approximate limit of performance for amplifying these features in different kinds of clinical or research samples and then sequencing with the available technologies of the time.

HIV-1 genomes are modeled differently depending on the reference accession, and whether the intent is to model the proviral DNA form or the viral mRNA form. The main reference genome used by the Los Alamos National Laboratory’s HIV Sequence Database is HXB2 GenBank: K03455.1, which was recently physically re-sequenced (Gener et al., 2021) and verified as a legitimate provirus with identical flanking LTRs. This sequence is important because all downstream analyses in the HIV Sequence Database are based either directly or indirectly on this historical species-specific reference.

With the advent of RNA-sequencing (Stark et al., 2019), it became convenient to represent the information contained in HIV differently. Today, the information contained in the HIV genome is now most often modeled as a full-length unspliced mRNA viral genome, with varying degrees of sequence annotation (e.g. RefSeq: NC_001802.1 gold-standard NCBI reference sequence; GenBank:MZ242719.1 with newer splice-associated open reading frames (GENERs) (Gener, 2022)). In this representation, the HIV LTRs were broken up at a leading small ∼100 bp R repeat (**Figure 2**). Multiple mapping is thus minimized for reads longer than 100 bp.

The main point of repeat sequence trimming is to remove problematic sequences while paying attention to limitations of mapping and variant calling in remaining repeat-associated (but ideally no longer repetitive) sequence. HIV as a system is an example where repeat sequence trimming (tail-trimming) has been adopted with minimal fuss.

### Sequencing and analysis considerations

In addition to considering the known biology of pathogens such as HIV or hMPXV, attention should be paid to the wet lab protocols used to generate sequencing data, and the planned analysis(es) which may depend on protocol used.

Reads can map to more than one place if there are repetitive sequences in a reference genome. Two relevant terms are secondary and supplemental alignments. These are described well in the documentation for a commonly used genome visualization tool the Integrative Genomics Viewer (Robinson et al., 2011), and are pasted here for clarity:

1. “A read may map ambiguously to multiple locations, e.g. due to repeats. Only one of the multiple read alignments is considered primary, and this decision may be arbitrary. All other alignments have the **secondary** alignment flag.” In this case, the entire read can map to multiple locations as in hMPXV ITRs. Source: https://software.broadinstitute.org/software/igv/book/export/html/6 on 01 September 2022.
2. “A chimeric alignment that is represented as a set of linear alignments that do not have large overlaps typically has one linear alignment that is considered the representative alignment. Others are called **supplementary** and have a supplementary alignment flag.” Source: https://software.broadinstitute.org/software/igv/book/export/html/6 on 01 September 2022.

Importantly, secondary/supplemental alignments need not be mutually exclusive. As repeats are difficult or impossible to unambiguously place (phase) given common short-read approaches, many genomic workflows include a repeat masking step to remove large low-complexity sequence from consideration while preserving assembly length. Repeat masking could solve the hMPXV ITR problem(s). However, this region is critical for current monkeypox-specific diagnostics. It also contains several protein-coding genes as mentioned above, including the allegedly most abundantly secreted OGP001 gene product. As such, it might benefit the global surveillance effort to monitor and track any changes in this region.

As for library considerations, it is important to consider the most common sequencing platforms available to most end-users in the sectors performing hMPX/hMPXV. Both long- and short-reads sequencing are currently used for most high-priority pathogens with large genomes. This includes hMPXV. Rather than focusing on which technology is better, different members of the community have used both to varying degrees during earlier surveillance during the 2022 outbreak (summarized in **Table 1**). Besides which platform(s) were used in each setting, earlier assemblies leverages both *de novo* and reference guided approaches. Competing priorities of quick assemblies with platform-specific errors were augmented with resequencing and either *de novo* short-read from reference-filtered data or hybrid assembly approaches.

**Table 1:**
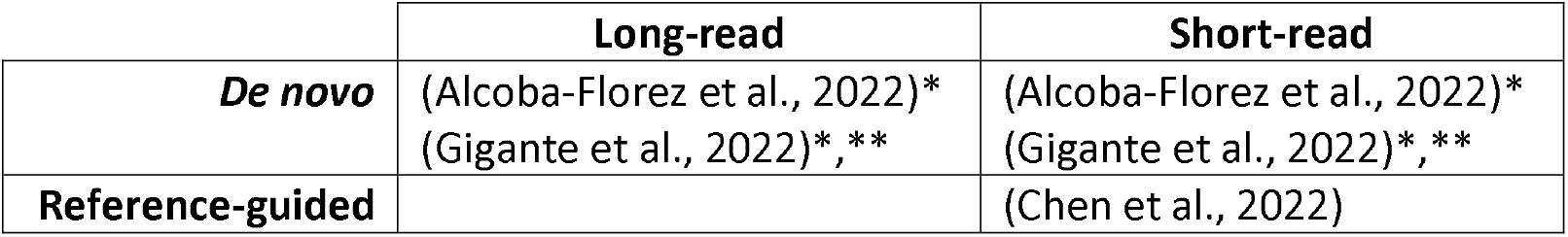
protocols/papers for hMPXV genome assembly. *Hybrid assembly approaches employ both long and short reads. **Reference hMPXV genomes (e.g., GenBank:MT903343) may be used to prioritize hMPXV-mapping reads for *de novo* assembly.

Regardless of assembly approach, resulting assemblies must be compared to available high-quality references to be able to call variants. Variant calling is a distinct process after either *de novo* assembly or consensus calling after mapping to a reference (*i.e*., reference-guided assembly). Because poxviruses have such low error rates, references were carefully made, and their quality assessed with orthogonal methods. For production-level (*i.e*., day-to-day) hMPXV assemblies, the accuracy of each assembly needs to be balanced with sequencing throughput to be able to adequately survey the number of cases seen during the present outbreak. In this context, because ILM sequencers are already present in many US public health labs already doing other routine pathogen surveillance, short-read reference-guided assembly is most used for hMPXV genomic surveillance. Other platforms such as ONT or Clear Labs (which uses ONT as part of a proprietary robotic and analytical solution) are also available in some public health and research settings across the local, state, and national level in the US.

### ITR problems and ITR tail-trimming proposal

The complete representation of terminal repeats has a time and a place. Where sequencing methods fall short, additional sequence can be distracting and cause problems (**Figure 3A-C**) that the current predominant public health workflows are not equipped to handle unambiguously. Problems relevant to hMPXV ITRs might include: resolving inter-ITR mapping (**Figure 3A**) and/or Intra-ITR mapping (**Figure 3B**). A third problem might occur (**Figure 3C**) when either ITR might appear to acquire a mutation, which then must be distinguished between its upstream or downstream counterpart. As a solution to the above problems, I propose hMPXV assembly ITR tail-trimming (tail-trimming for short; **Figure 3D**), which has 2 parts. Firstly, trimming (removing or deleting) the sequence downstream of the first ∼300 bp of the 3’ LTR (right green arrow). Secondly, annotating the medial ∼300 bp of the 5’ ITR and remaining (also medial) 3’ ITR as phasable, while annotating the tail or distal segment of the of the 5’ ITR as conditionally phasable, where read length and sequencing depth determine phasability. See “**Data availability**” section below for specifics.

**Figure 3:**
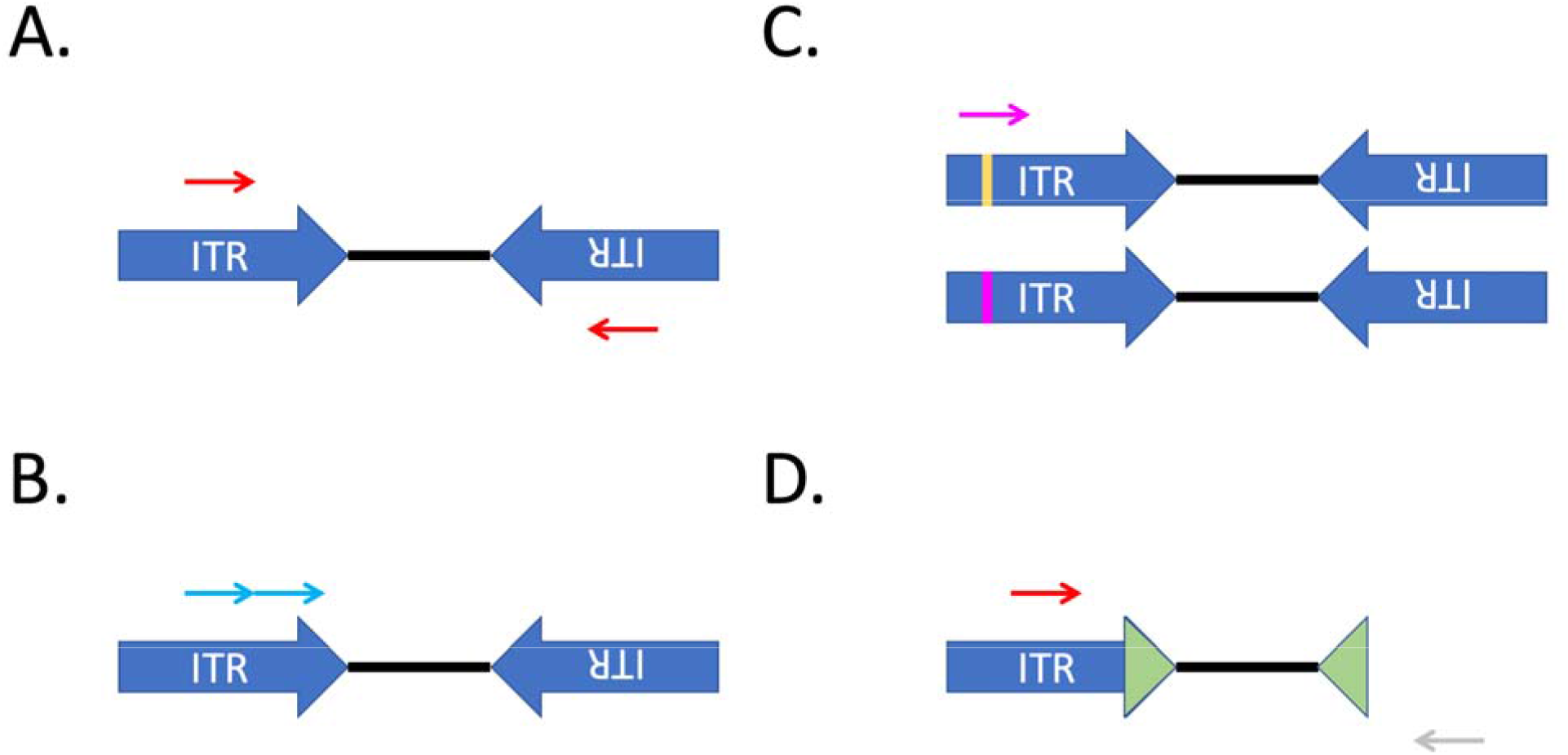
Problems inherent to hMPXV ITR sequence analysis, and a possible solution. Inter-ITR mapping. Reads (thin red arrow) from ITR sequence (large blue arrow) cannot be phased unless they are anchored to non-ITR sequence. This can happen with 1.) reads longer than either ITR or 2.) reads with downstream/upstream non-ITR sequence with bridging contiguous sequence running into the ITR. Neither 1 or 2 are shown to emphasize the current state of most hMPXV public health sequencing. Red arrows denote secondary alignments. Note that the absolute orientation of arrows are arbitrary. For example, NCBI displays them as <-…->. This is likely because most of the open reading frames in the ITR occur in the direction of the arrow. An important relationship of the ITR sequences is that each ITR is inverted relative to the other. Reads/ITR not show to scale. **3B**: *Intra-ITR mapping*. This was not an issue after kmer analysis if using PE150 and insert size ∼300 bp (negative results not shown). **3C**: Either ITR might inherit a mutation, which then must be distinguished between its upstream or downstream counterpart. Repeat DNA extraction and resequencing can help to resolve allelic artifactual heterogeneity. Note that most genomic surveillance performed in public health settings is done as single runs. Technical duplicates are not routinely done in public health labs. Higher capacity labs may reextract and resequence samples for quality. **3D**: I propose hMPXV assembly ITR tail-trimming (tail-trimming for short), which has 2 parts. Firstly, trimming (removing or deleting) the sequence downstream of the first ∼300 bp of the 3’ LTR (right green arrow). Secondly, annotating the medial ∼300 bp of the 5’ ITR and remaining (also medial) 3’ ITR as phasable, while annotating the tail or distal segment of the of the 5’ ITR as conditionally phasable, where read length and sequencing depth determine phasability. See “Data availability” section below for specifics.

As a proof-of-principle, I took reads from a public hMPXV sample (BioSample:SAMN30416950; run accession SRR21143274) collected on 25 July 2022 and recently sequenced (submitted to SRA on 19 August 2022) and mapped these to NC_063383.1 (untrimmed) and NC_063383.1_tt (ITR tail-trimmed) assemblies (**Figure 4**).

**Figure 4:**
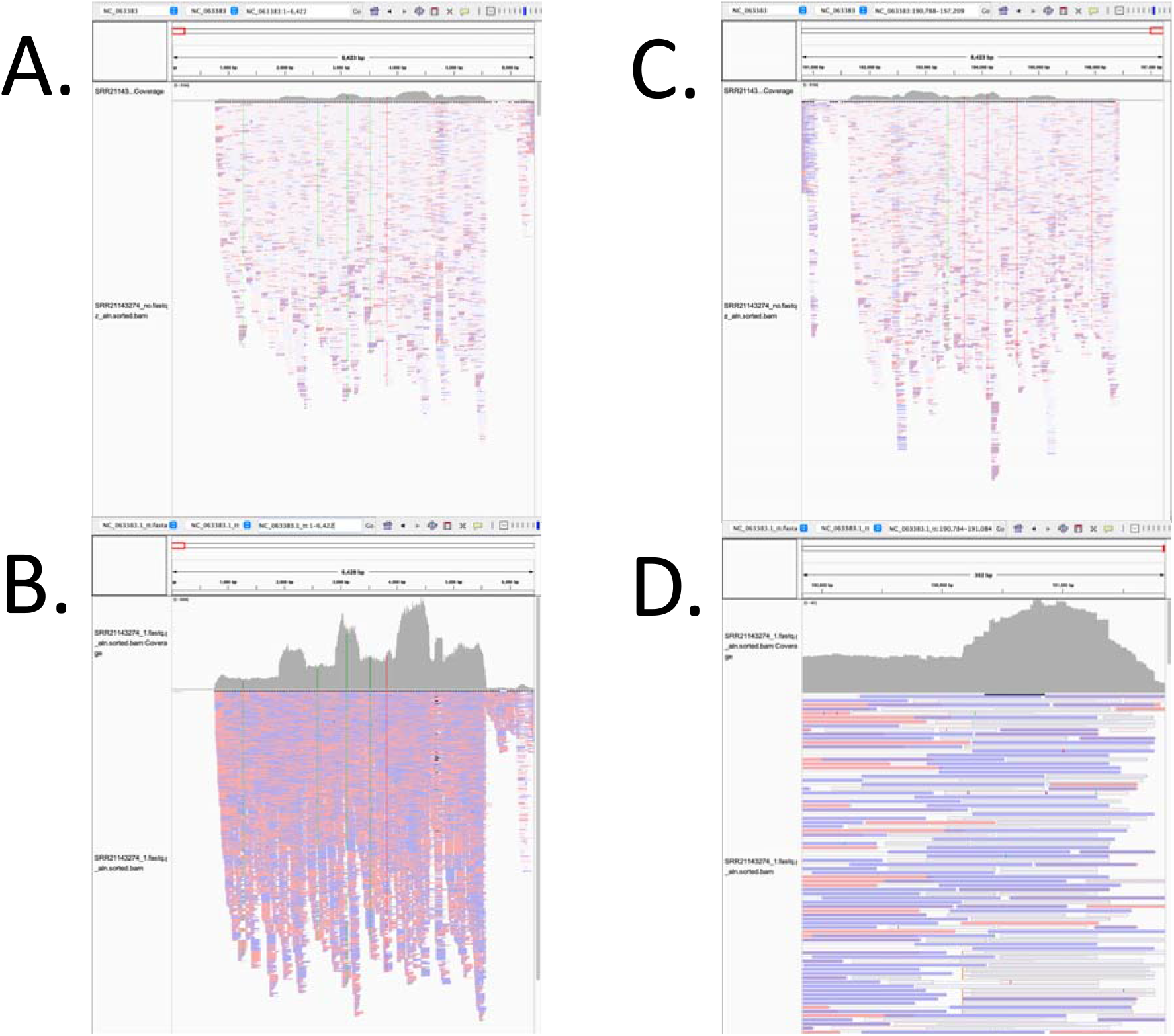
ITR tail-trimming mitigates secondary alignments. The following MPXV WGS sequencing data was used as an example due to its high coverage over the ITRs: CA-LACPHL-M10162_081822_5x_01 of BioSample:SAMN30416950; run accession SRR21143274. This was submitted by Los Angeles County Public Health Lab microbial pathogen submission group (LACPHL) under BioProject:PRJNA864832. Minimap2 (Li, 2018) was used to remap SRR21143274 reads to NC_063383.1 or to a tail-trimmed NC_063383.1 “NC_063383.1_tt.fasta.” Samtools (Wysoker et al., 2009) was used to prepare for visualization in IGV. Windows shown correspond to ITR segments only. Reads are colored based on their orientation: salmon = forward; violet = reverse. Single nucleotide polymorphisms are represented by green and red vertical stripes. **4A**: SRR21143274 remapped to NC_063383.1 5’ ITR. Note the abundant pale reads. **4B**: SRR21143274 remapped to NC_063383.1 3’ ITR. Note the abundant pale reads. **4C**: SRR21143274 mapped to NC_063383.1_tt 5’ ITR. Squished. Coverage is lower near the downstream 5’ ITR. **4D:** SRR21143274 mapped to NC_063383.1_tt 3’ ITR. Collapsed. Several reads mapped to upstream of the window. Supplemental alignments are minimized. Note that the range of the phasable segments can be extended depending on the insert size and/or platforms (e.g., average read length). Note that variants identified in are not within the phasable segments of the ITR.

For this sample, ITR tail-trimming was able to mitigate most of the secondary mapping. Recalling that ITR reads and any variants distal to the proximal ∼300 bp would not be able to be phased. However, they could at least be displayed together and analyzed separately from the phasable hMPXV genome.

Tail-trimmed reference assemblies (**Figure 2B, Figure 3D, Figure 4**) would better meet the current needs of all amplicon-based assembly and variant calling workflows for hMPXV. Because this is an analytical method, ITR tail-trimming would not disrupt current wet lab protocols. As such, it approximates *gratis*. ITR tail-trimming facilitates ITR inspection and ergo tracking ITR change over time which might impact current hMPX-specific diagnostics. This hack may be extensible to other poxviruses and to other viruses with terminal repeats that are longer than the reads used to interrogate sequenced samples.

Besides manual inspection, community resources may leverage this method to follow variability in previously underappreciated ITRs. A major open sequence analysis webserver caller Nextclade (Aksamentov et al., 2021) does not currently explicitly include non-coding sequences in their gene-specific dropdowns. This is in part because this tool is not meant to be an explicit genome viewer (as opposed to IGV, the US National Center for Biotechnology Information (NCBI)’s Graphics view of the reference genome nucleotide records, UCSC genome browser (which pulls from the RefSeq:NC_063383.1), others). Until ITR tail-trimming is adopted widely and until noncoding regions are included explicitly by Nextclade (if curators choose to include them), Nextclade does display OPG001, OPG002, OPG003, and OPG015, which serves as a proxy to direct ITR observation. However, because these genes occur on repeat elements, their variant calling and Nextclade flags are subject to misinterpretation. This is not obvious unless one is aware of the nature of repeats generally (as above) and unless one takes the time to appreciate these elements and analyze them appropriately. This is mentioned without judgement, but with encouragement to redress the issue for public health community members studying this emerging global pathogen. With ITR tail-trimming and segment annotation, these regions and open reading frames can be effectively flagged and subsequently handled appropriately as diploid depending on the methods used to generate reads. Deviation from reference(s) can be more easily appreciated and followed if/when it occurs.

Implementing hMPXV genomic tail-trimming by treating diploid ITRs as conceptually distinct from single-copy haploid non-ITR hMPXV sequence may help focus surveillance efforts at these problematic regions. Importantly, ITR tail-trimming does not preclude using both full-length 5’ and 3’ ITRs when reads are long enough to extend the phasable sequence (green arrows in **Figure 3D**). In the time that it takes for longer-read sequencing and/or metagenomic sequencing to be adopted, methods to improve interpretation and limitations of sequence analysis may aid in the ongoing hMPX/hMPXV genomic surveillance effort.

## Data Availability

All data produced in the present work are contained in the manuscript.

## Data availability

The phasable segments of the hMPXV include the following sequences which occur twice (once at the 5’ ITR and then the reverse-complement toward the 3’ ITR; 2x total) in the current implementation of NC_063383.1:

~~~
>phasable_ITR_segment_ NC_063383.1
GCTCATCGACAGCCATGAAATCTACCGACTCCATGGTGCGAATCGCACTGTCTTATTCGCCATTGATTTTCATTTTTTATAATTATGTACATGTTTTCCTTCTATTCTCAAGAGTCTACAAAAATATATTTTTTCGATATCTAAGTACTAAGTTTTTTTACTGTTTTTGTTACTGTCTTCCATTCTTCTAACTAAAGATCTGAGATAAATTATACAATCTTCGCTATCGAACCATTTTTGTAGTCTAAAGCCTGAAGTAATTAACCAACTGTTTTTATTAGTGGCTTTTTTCGATCTATC
//
~~~

I propose annotating the upstream 1..6122 (base-1; inclusive) of the 5’ ITR as the conditionally phasable segment, and tail-trimming the terminal 6122 bases of the 3’ ITR to simplify genomic analyses. This assumes PE 150, and that reads longer than average insert of 300 should help phase reads to their respective poles.

## Funding

This work was supported by Cooperative Agreement Number NU60OE000104-02, funded by the Centers for Disease Control and Prevention through the Association of Public Health Laboratories. Its contents are solely the responsibility of the author and do not necessarily represent the official views of the Centers for Disease Control and Prevention or the Association of Public Health Laboratories.

## Conflict of interest statement

I have received travel support in the form of poster bursaries from Oxford Nanopore Technologies, Oxford, UK. I am on the editorial board of AIDS.

## Acknowledgements

I would like to sincerely thank the curators in NCBI, Nextstrain, and other pathogen sequence databases who have maintained their respective databases. I would also like to thank members of the SPHERES and Staph-B communities for their invaluable discussions. It takes a village.

## Notes

### Author Declarations

Example anonymized and dehosted sequencing data was downloaded from the US National Center for Biotechnology Information (NCBI) Short Read Archive (SRA). The following MPXV WGS sequencing data was used as an example due to its high coverage over the ITRs: CA-LACPHL-M10162_081822_5x_01 of BioSample:SAMN30416950; run accession SRR21143274. This was submitted by Los Angeles County Public Health Laboratories microbial pathogen submission group (LACPHL) under BioProject:PRJNA864832. The hMPXV Clade IIb reference genome used was RefSeq:NC_063383.1 which was used to derive the tail-trimmed assembly mentioned in the text with documentation to make.

